# Within-country heterogeneity in patterns of social contact relevant for tuberculosis infection transmission, prevention, and care

**DOI:** 10.1101/2025.01.21.25320893

**Authors:** Kate E. LeGrand, Anita Edwards, Mbali Mohlamonyane, Njabulo Dayi, Stephen Olivier, Dickman Gareta, Robin Wood, Alison D. Grant, Richard G. White, Keren Middelkoop, Palwasha Khan, Nicky McCreesh

## Abstract

*Mycobacterium tuberculosis* (*Mtb*) transmission is driven by variable social, environmental, and biological factors, including the number and duration of indoor contacts. Social contact data can provide information on potential transmission patterns, but is underutilised outside the field of mathematical modelling. We explore three contexts where contact data can provide valuable insights: 1) household contact tracing; 2) infection prevention and control measures (IPC) in congregate settings; and 3) contamination in cluster randomised trials (CRTs).

A social contact survey was conducted in three communities with comparable population sizes in South Africa: an urban township and peri-urban and rural clinic catchment areas. Participants reported congregate settings visited over 24-hours, visit durations, and estimated numbers of people present. To correspond with the three contexts, we estimated the proportion of contact hours occurring 1) within the home; 2) in congregate settings outside the home; and 3) outside the participants’ communities.

Participants reported a mean of 27.0 (rural), 55.2 (peri-urban), and 73.0 (urban) contact hours. The proportion within the home was similar among rural and peri-urban participants (76.8% and 71.7%), compared to urban (48.6%). Congregate settings visited varied; urban participants spent the most contact hours in retail/office settings (19.9%), peri-urban participants in community-service buildings (20.4%), and rural participants in other peoples’ homes (25.5%). Urban participants reported the highest proportion of contact outside the community (67.0%) compared to rural (38.8%) and peri-urban (21.5%) participants.

The heterogeneity in contact patterns has implications for TB interventions. Household contact tracing may be most effective in the rural community where household contact was highest. The diverse range of congregate settings visited suggests that prioritising IPC measures in these locations may enhance their overall efficacy. Considering contact patterns when designing clusters may reduce contamination risk in CRTs. Tailored interventions, informed by local contexts, are essential to reduce TB burden.

## Introduction

Tuberculosis (TB) is a pervasive threat to global public health. In 2023, an estimated 10.8 million developed TB, and an estimated 1.25 million people died from the disease.(1) South Africa contends with a particularly high TB burden with a person developing incident TB every two minutes, while every nine minutes the disease claims another life.(1,2) Reducing TB burden requires disrupting the transmission of *Mycobacterium tuberculosis* (*Mtb*), the bacillus that causes the majority of TB disease.

*Mtb* spreads from person to person through airborne transmission, influenced by highly variable social, environmental, and biological factors, including the frequency and duration of social contact, as well as the time spent in poorly ventilated spaces.(3) While sustained household exposure has historically been considered the primary route for transmission, recent molecular and epidemiological evidence suggests that more than 80% of *Mtb* transmission in high burden settings is attributable to exposure outside the household.(4–7) Identifying congregate settings where transmission may occur requires a comprehensive understanding of contact patterns within the broader population. Such investigations are pivotal for elucidating the dynamics of *Mtb* transmission and developing effective strategies.

The incubation period from the acquisition of *Mtb* infection to the onset of TB disease poses a challenge for intervention implementation. Approximately 5% of healthy adults who acquire Mtb infection will progress to TB disease within the first two years, while others develop the disease a decade or longer after exposure.(8,9) This latency makes it difficult to identify the propitious conditions for transmission. Social contact surveys can partly address this limitation by capturing detailed information on social behaviours and population movement. These surveys typically inquire about close contacts (face-to-face conversations or physical touch).(10,11) While less common, some surveys also include data on casual contacts (people ‘sharing air’ in indoor settings), which may be more relevant for airborne infections like *Mtb*.(12,13) While social contact data offer valuable insights, contact patterns can differ systematically across settings, which has implications for understanding the dynamics of transmission.(10,14,15)

Given the heterogeneity of contact patterns across populations, a uniform approach for interventions may be insufficient to interrupt transmission. To bridge the knowledge gap between social contact patterns and their implications for TB interventions, we analysed data from a social contact survey conducted in three geographically distinct communities in South Africa. We examined three TB contexts where social contact data can provide valuable insights: Context 1) Household contact tracing, where we estimate the proportion of contact hours occurring within participants’ own homes; Context 2) Infection prevention and control strategies, where we estimate the proportion of contact hours occurring in various congregate settings; and Context 3) Contamination in cluster randomised trials, where we estimate the proportion of contact hours occurring outside the community. For this third context, we considered the communities as clusters in a trial, and contact outside the community as a measure of mobility. Contamination in CRTs, resulting from mobility between clusters and the wider population, can dilute the intervention impact and lead to an underestimation of the true effect.(16)

## Materials and methods

### Study communities

In 2019, a social contact survey was conducted in three communities in South Africa with comparable population sizes: an urban township in Western Cape (WC) province and peri-urban and rural areas in KwaZulu-Natal (KZN) province (Figure 1).(17) High TB notification rates and HIV prevalence dominate the public health landscape of the three study communities, yet strong collaborations facilitate research and programmes for care and prevention.(18–20)

**Figure 1.**
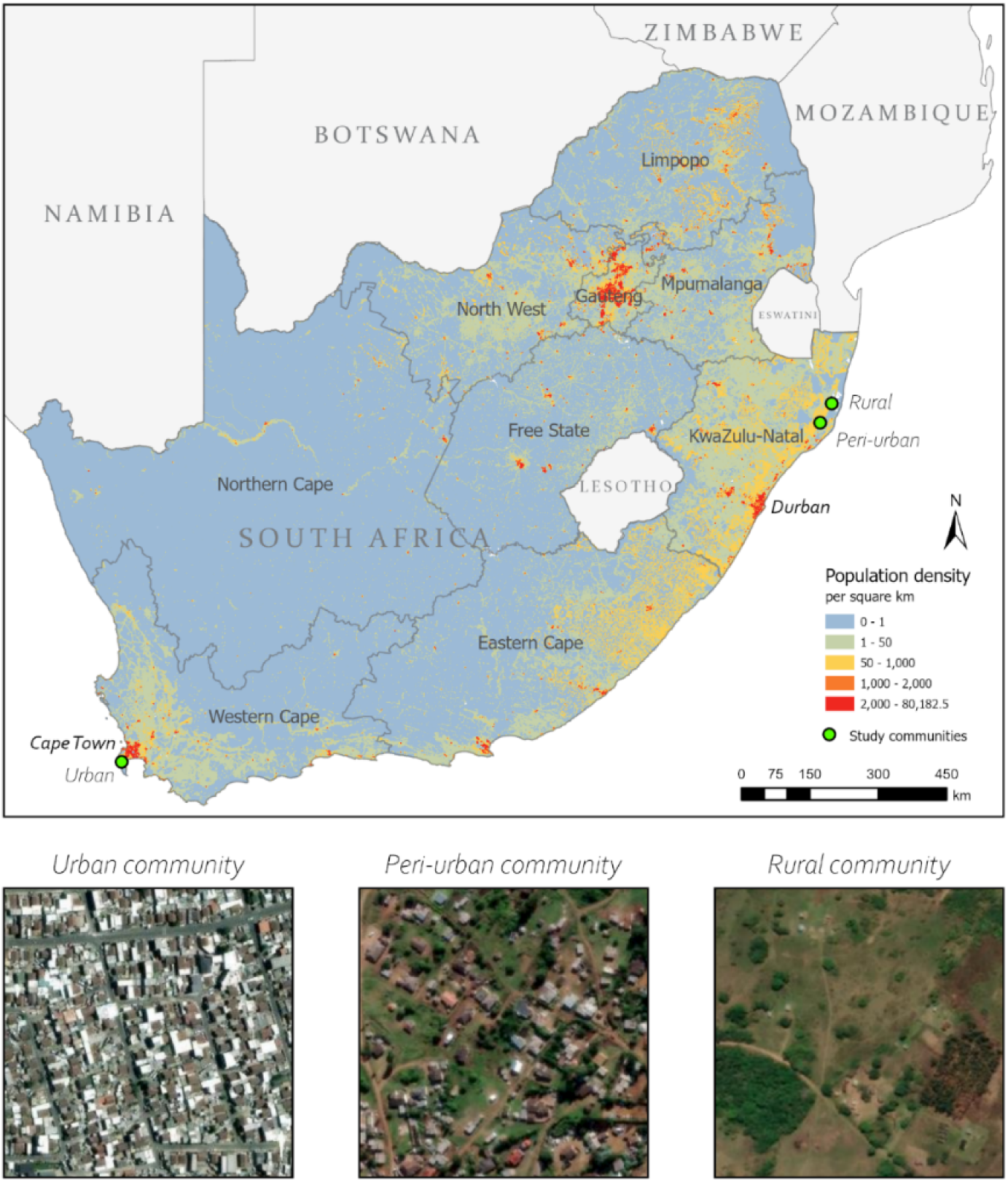
Map of study community locations and population densities in South Africa, 2020. km = kilometre. Area of each satellite imagery inset(22) is approximately 0.1 km^2^. National population density from WorldPop.(23)

The urban community is an established research site with biennial censuses located approximately 40 kilometres (km) south of Cape Town. This well-demarcated township has an area of about 0.5 km^2^ and a population of 27,000, making it the most densely populated community in this study (54,000 people/km^2^).(17) The community comprises a formal sector with numbered housing plots with basic utilities, and an informal sector with crowded shack dwellings and limited communal services, which are characteristic of low socioeconomic communities.(21) The weighted average household size among survey participants from the urban community is 3.9 people.

The peri-urban and rural communities are each a catchment area of a primary healthcare clinic within a health and demographic surveillance area. In the overall surveillance area, most households have access to basic utilities with over 95% of households having electricity and toilets, and 66% have access to piped water. At their closest point, the peri-urban and rural communities are approximately 12 kilometres apart. The peri-urban community spans an area of about 165 km^2^ with a population of 25,000 (population density of 152 people/km^2^). The rural community has the lowest population density of the three communities (85 people/km^2^) with a population of approximately 22,000 and an area of 260 km^2^. Both communities have large household sizes, with peri-urban and rural participants reporting a weighted average of 15.2 and 10.9 people, respectively.

### Survey design and data collection

In the urban community (WC), 1,530 adults aged 15 and older were randomly sampled by age and sex from a population of approximately 27,000 adults. In the peri-urban and rural communities (KZN), 3,093 adults ages 18 and older were randomly sampled by residential area from a total population of about 46,000 adults. For this comparative analysis, we excluded respondents from WC aged 15-18, as the KZN survey on collected data from people aged 18+ and older only. The methods for survey design and data collection have been previously described in full.(17)

Participants were interviewed in-person and asked to recall their activities on a randomly assigned day in the past week in the KZN communities, and on the day preceding the interview in the WC community. Participants listed all buildings visited, and for each building the type of building, the location of the building, the duration of the visit, and the estimated number of people present halfway through the visit. Additionally, participants reported information about their use of transportation, including the vehicle type, duration, and number of people present at the start of their journey. Information on sociodemographic strata, including age, sex, and employment status was also collected. Details on the survey questions analysed for this study are available in the Appendix.

### Statistical analysis

We analysed the social contact survey data to assess the potential implications of contact patterns on the effectiveness and evaluation of tuberculosis interventions. We classified the location of contact as relevant for three TB contexts: Context 1) Household contact tracing, where we estimate the proportion of contact time occurring within participants’ homes; Context 2) Infection prevention and control settings, where we estimate the proportion of contact time occurring in different congregate settings; and Context 3) Contamination in cluster randomised trials, where we estimate the proportion of contact time occurring outside the community as a measure of mobility. The WC survey asked whether the building visited was located within the community, whereas the KZN survey asked participants to name the administrative unit within which the building was located. The locations of the KZN buildings were validated using ESRI ArcMap (Version 10.8.1) (Appendix). Data cleaning, analysis, and visualisation of results were carried out in R.

For airborne infections like *Mtb* where transmission can occur in shared air without close contact, reporting the number of close contacts (i.e. people spoken to or touched) alone does not capture the total amount of exposure an individual has to others. To address this limitation, we estimated cumulative indoor contact hours as the product of the duration of each building visit or a transport journey and the number of people present. We set a maximum of 20 people in any building type, and 10 for private cars, as it is unlikely that a participant could have sufficient contact with every adult above this value to allow infection transmission. To account for the potential effects of larger group sizes, we conducted sensitivity analyses with caps of 50 and 100 people in buildings. (Appendix Table S3). Contact hours were weighted for each community according to the study population composition by age group (18-24, 25-44, 45+) and sex. To account for the differences in the proportions of participants who recalled their activities on each day of the week or weekends, contact hours were also weighted by day of the week. Missing values in the number of people present were imputed by averaging the value from the same conditions (i.e., same community and type of building visited or transport used).

We estimated the number of indoor contact hours for each location setting by community and sociodemographic strata (sex, age, group, and employment status). To understand average contact trends, we calculated the mean number of indoor contact hours (MICH) by taking the sum of contact hours reported for each stratum and dividing it by the total number of participants from that stratum. We then calculated the proportion of contact hours that occurred within the home, outside the home by congregate setting, and outside the community. We generated 95% plausible ranges of MICH and proportions by bootstrapping, during which, missing values were imputed by selecting a random value given the same conditions (Appendix).

While absolute mean contact hours will provide some indication of the risk of transmission by location in each community, differences in TB prevalence between communities will influence transmission risk, making direct comparisons between communities challenging. We therefore report proportions of contact hours to facilitate comparisons. For example, a higher proportion of contact hours in municipal buildings one community would suggest that targeted IPC measures in these buildings could result in a greater relative reduction in community-wide TB incidence in that community. By comparing the proportions of contact hours across communities, we can identify interventions that may be most contextually relevant and impactful.

The impact of household contact tracing on *Mtb* transmission in the wider population may be limited, and may vary between settings.(24) In Context 1, to assess the potential for household contact tracing, we compared the proportion of contact hours occurring within and outside a participant’s own home.

Contact patterns outside the household are particularly relevant in understanding how TB spreads, and are useful for informing the implementation of IPC interventions in congregate settings.(25) In Context 2, we examined the proportion of contact hours spent outside the home in ten groupings of buildings and transport: community services (e.g., clinics, hospitals, churches, libraries), food and leisure, retail and office, schools, workshops (e.g., mechanic shops, factories), own homes, other homes, other buildings, unknown buildings, and an overall transport category. A full list of these groupings is available in the Appendix (Table S1).

Understanding the geographic location of where contact occurs can have an effect on the design and evaluation of CRTs.(26) Contamination due to mobility between clusters and the wider population can dilute the impact of interventions, leading to an underestimation of their true effect.(16) In Context 3, we considered the communities in this study as clusters and analysed the proportion of contact hours spent outside the community as a measure of mobility, which can be an indicator of contamination. As it was not possible to definitively determine whether contacts made during transport were with individuals from within or outside the community, contact hours in transport were excluded from the proportion calculations in Context 3.

## Results

### Sociodemographic characteristics of survey participants

A summary of the included participants is provided in Table 1. A total of 4,623 individuals were sampled across the three study communities resulting in 2,673 (58%) participants in the analysis. Of the 1,530 individuals sampled in Western Cape, 1,115 (73%) completed an interview, of whom, 52 (5%) were under 18 years old and excluded, resulting in 970 (87%) for analysis. Of the 3,093 individuals sampled in KwaZulu-Natal, 1,703 (55%) completed an interview, of whom 842 (49%) and 861 (51%) were from the peri-urban and rural communities, respectively.

**Table 1.** Sociodemographic characteristics of survey participants.

| Participant characteristics | Urban |  | Peri-urban |  | Rural |  |
| --- | --- | --- | --- | --- | --- | --- |
|  | Count | Percentage | Count | Percentage | Count | Percentage |
| **Sex** |  |  |  |  |  |  |
| Female | 481 | 49.6 | 466 | 55.3 | 485 | 56.3 |
| Male | 489 | 50.4 | 376 | 44.7 | 376 | 43.7 |
| **Age group** |  |  |  |  |  |  |
| 18-19 | 76 | 7.84 | 63 | 7.48 | 55 | 6.39 |
| 20-29 | 376 | 38.8 | 244 | 29.0 | 251 | 29.2 |
| 30-39 | 333 | 34.3 | 152 | 18.1 | 155 | 18.0 |
| 40-49 | 133 | 13.7 | 123 | 14.6 | 104 | 12.1 |
| 50+ | 52 | 5.36 | 260 | 30.9 | 296 | 34.4 |
| **Time lived in house** |  |  |  |  |  |  |
| Less than 2 years | 224 | 23.1 | 20 | 2.38 | 45 | 5.23 |
| 2-5 years | 234 | 24.1 | 76 | 9.03 | 21 | 2.44 |
| More than 5 years | 511 | 52.7 | 746 | 88.6 | 795 | 92.3 |
| Unknown | 1 | 0.103 | 0 | 0 | 0 | 0 |
| **Time lived in community** |  |  |  |  |  |  |
| Less than 2 years | 85 | 8.76 | 9 | 1.07 | 4 | 0.465 |
| 2-5 years | 166 | 17.1 | 38 | 4.51 | 19 | 2.21 |
| More than 5 years | 718 | 74.0 | 795 | 94.4 | 838 | 97.3 |
| Unknown | 1 | 0.103 | 0 | 0 | 0 | 0 |
| **Employment status** |  |  |  |  |  |  |
| Full-time | 379 | 39.1 | 237 | 28.1 | 91 | 10.6 |
| Part-time or casual | 191 | 19.7 | 33 | 3.92 | 35 | 4.07 |
| Not employed | 393 | 40.5 | 452 | 53.7 | 644 | 74.8 |
| Unknown | 7 | 0.722 | 120 | 14.3 | 91 | 10.6 |
| **Day reported** |  |  |  |  |  |  |
| Monday | 173 | 17.8 | 106 | 12.6 | 133 | 15.4 |
| Tuesday | 180 | 18.6 | 130 | 15.4 | 112 | 13.0 |
| Wednesday | 166 | 17.1 | 117 | 13.9 | 122 | 14.2 |
| Thursday | 119 | 12.3 | 139 | 16.5 | 111 | 12.9 |
| Friday | 70 | 7.22 | 124 | 14.7 | 137 | 15.9 |
| Saturday | 83 | 8.56 | 119 | 14.1 | 126 | 14.6 |
| Sunday | 179 | 18.5 | 107 | 12.7 | 120 | 13.9 |
| **Total participants** | 970 |  | 842 |  | 861 |  |

In the urban community, participants were younger with 80.9% (785/970) under age 40, compared to 54.5% (459/842) in the peri-urban and 53.5% (461/861) in the rural communities, reflecting the demographics of the study populations (Appendix Table S2). High unemployment was reported by all three communities; urban participants reported the lowest proportion at 40.5% (393/970), compared to nearly three quarters of rural participants (74.8%, 644/861), and more than half of peri-urban participants (53.7, 452/842). Nearly all peri-urban (94.4%, 795/842) and rural (97.3%, 838/861) participants were long-term residents of their communities (five or more years), compared to 74.0% (718/970) of urban participants.

### Mean indoor contact hours

Our analysis of social contact data across the three communities revealed distinct contact patterns overall and by sociodemographic strata (Figure 2; Appendix Table S2). Considering all congregate settings, participants from the rural community reported the highest MICH of 73.0 hours (95% CI: 69.3-76.9). This was followed by peri-urban participants, who reported a MICH of 55.2 (CI: 51.7-57.7). Urban participants reported the lowest MICH, 27.0 (CI: 25.4-28.4). Rural participants generally reported the highest MICH across the strata, with the highest among ages 45 and older (81.0, CI: 72.0-90.4). Participants from the urban community reported the lowest MICH across all strata, with part-time employees showing the lowest (24.6, CI: 20.2-27.3)

**Figure 2.**
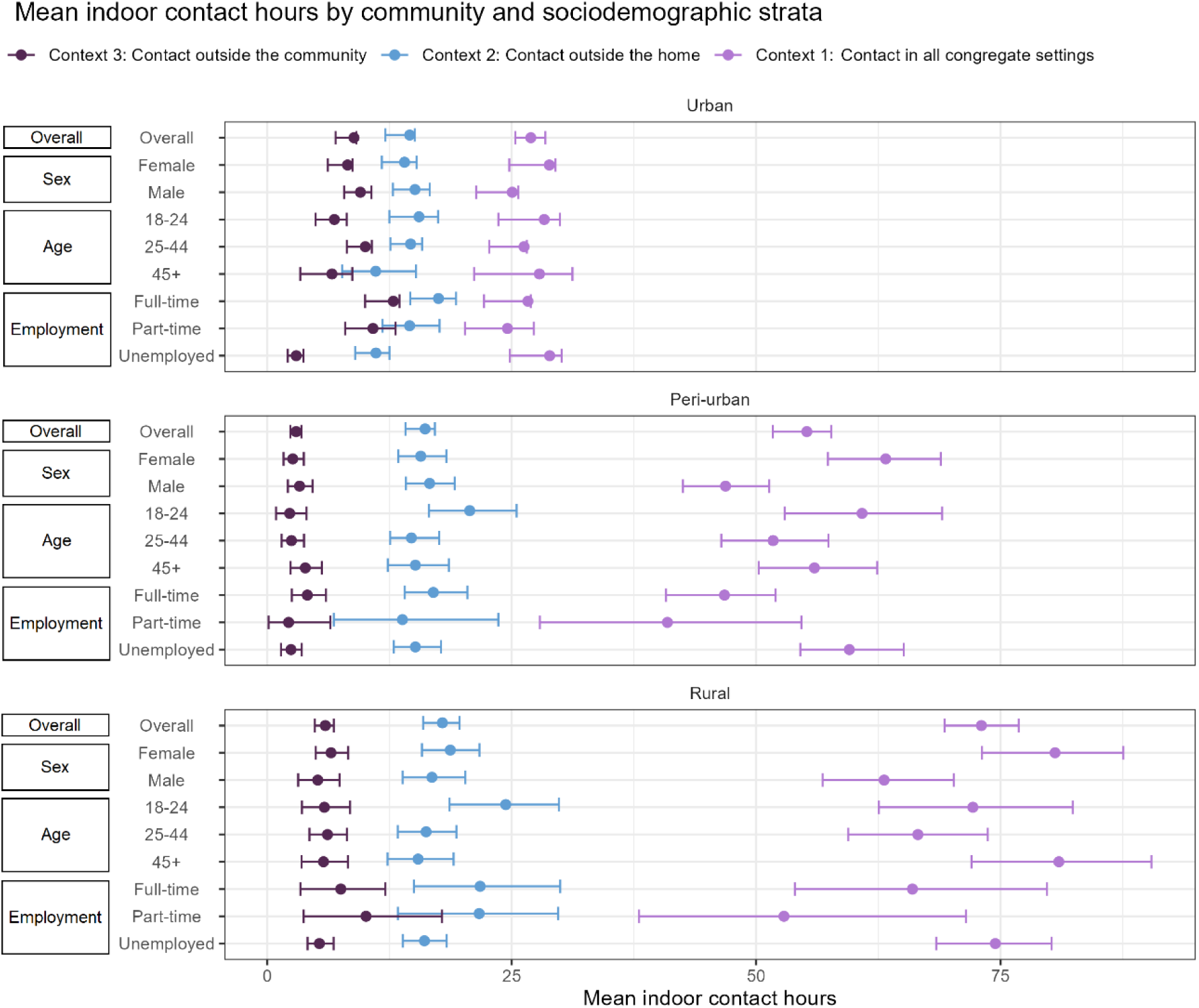
Mean indoor contact hours reported by community and sociodemographic strata. Contact in all congregate settings includes own home and transport; Contact outside the home includes transport; Contact outside the community excludes transport.

Contact patterns in congregate settings outside the home were more consistent across the communities. Rural participants had the highest MICH, 17.9 (CI: 15.9-19.7). Peri-urban was similar, with participants reporting a mean of 16.1 (CI: 14.1-17.2), and urban participants had the lowest MICH (14.6, CI: 12.1-15.1). These trends extended to the strata, where rural participants generally reported higher means than the other two communities. Rural participants ages 18-24 reported the highest MICH (24.4, CI: 18.7-29.8). Additionally, full-time rural employees reported the next highest MICH (21.8, CI: 15.0-30.0), surpassing those from the urban (17.5, CI: 14.6-19.3) and peri-urban (17.0, CI: 14.1-20.5) communities.

Disparities in contact patterns emerged when examining contact occurring exclusively outside the community. Urban participants reported the highest MICH (8.9, CI: 7.0-9.1), followed by rural participants (5.9, CI: 4.9-6.8), and peri-urban participants (3.0, CI: 2.4-3.5). Regarding the strata, full-time (12.9, CI: 10.0-13.5) and part-time (10.8; CI: 8.0-13.1) employees from the urban community reported the highest MICH. There were slight differences in MICH by sex, with males having higher means than females in urban and peri-urban settings. Urban males had a MICH of 9.5 (CI: 7.9-10.7) compared to females with 8.2 (CI: 6.2-8.7). In the peri-urban community, these values were 3.3 (CI: 2.1-4.6) and 2.6 (CI: 1.7-3.7), respectively. Rural females reported a MICH of 6.5 (CI: 5.0-8.3) compared to males with a mean of 5.2 (CI: 3.2-7.4).

#### Context 1: Household contact tracing

Participants across the three communities generally spent more contact hours in their own homes then outside the home (Figure 3A). However, there were notable differences in these proportions. Rural participants had the highest proportion of household contact hours (76.2%, CI: 73.5%-79.0%), followed by the peri-urban participants (70.8%, CI: 67.6%-73.8%). By contrast, urban participants spent less than half of contact hours in the household (46.0%, CI: 42.9%-49.0%). Contact hours outside the home were highest in the urban community across all sociodemographic strata, with full-time employees having the greatest proportion (65.8%, CI: 61.7%-69.8%).

**Figure 3.**
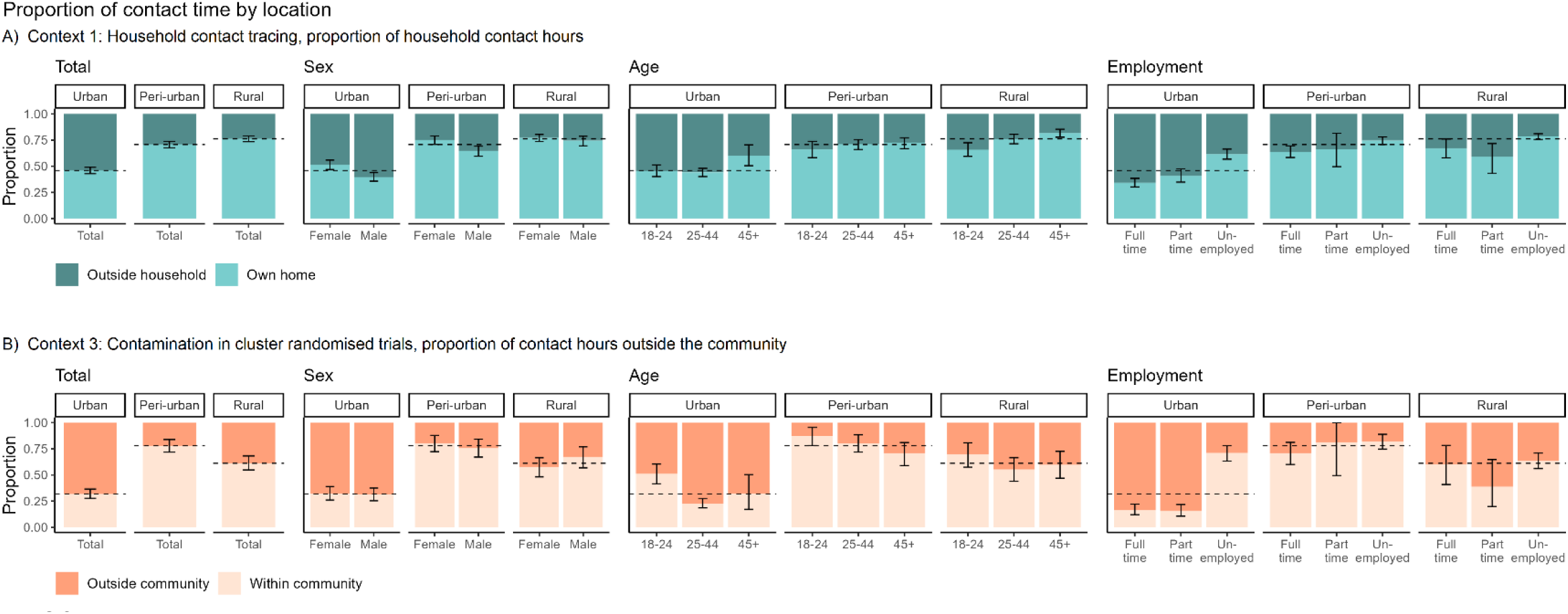
Proportion of contact hours occurring inside and outside households and communities. Dashed lines indicate the overall mean proportion in each community. A) All building types and transport were included for congregate settings outside the household. B) Transport was excluded from contact occurring inside and outside the community.

#### Context 2: Infection prevention and control strategies

The proportion of contact hours spent in specific congregate settings outside the home varied substantially across the urban, peri-urban, and rural communities (Figure 4; Appendix Table S6). Urban participants had a more diverse range of activities and interactions outside their homes; retail and office buildings accounted for the largest proportion of contact hours (19.2%, CI: 13.8%-21.8%), followed by food and leisure settings (15.7%, CI: 12.8-20.3%). Other people’s homes, community services, and schools, each accounted for less than 10% of contact hours from urban participants. By contrast, the highest proportion of contact hours among rural participants was within other people’s homes (25.1%, CI: 20.1%-30.5%) and in school (21.4%, CI: 16.3-27.4%). In peri-urban areas the greatest proportion of contact hours occurred in buildings providing community services (19.7%, CI: 15.7%-24.5%), followed by time in other people’s homes (16.1%, CI: 12.3%-20.5%). Contact hours reported in transport were relatively similar across communities, with urban participants reporting the largest proportion (11.7%, CI: 9.9%-15.0%), closely followed by peri-urban (11.2%, CI: 9.1%-13.7%) and rural (10.4%, CI: 8.3%-13.0%) participants.

**Figure 4.**
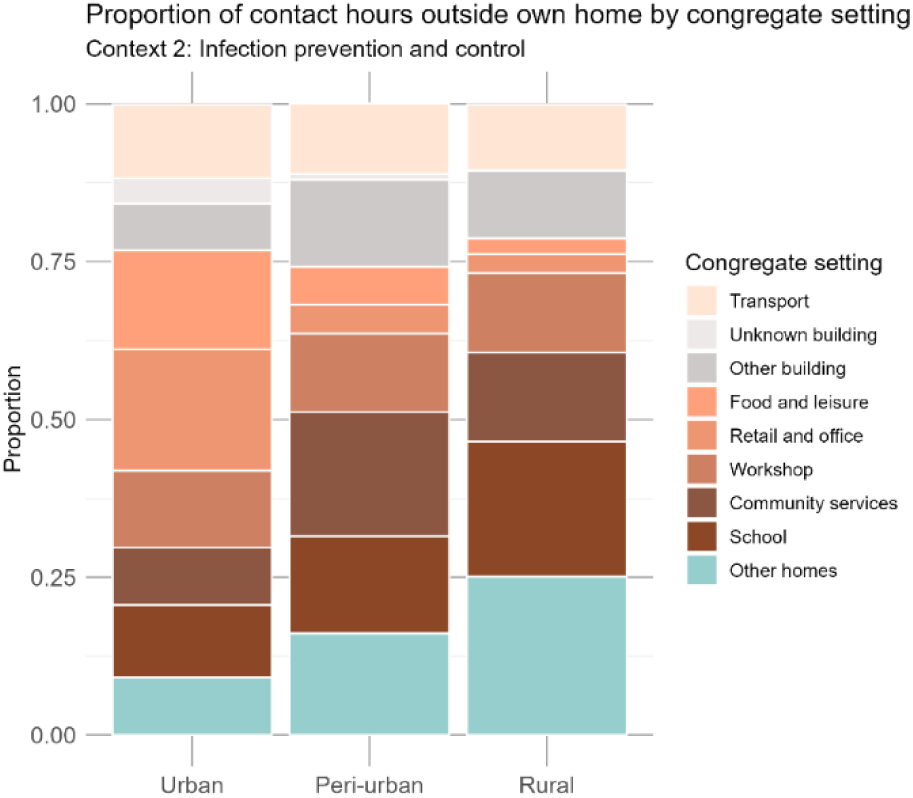
Proportion of indoor contact hours occurring outside the household attributable to each congregate setting.

#### Context 3: Contamination in cluster randomised trials

The differences in contact patterns were more pronounced when examining the proportion of contact hours spent within and outside the communities (Figure 3). Urban participants reported the highest proportion of contact hours outside the community by far, reaching 68.2% (CI: 63.5%-72.4%). This finding stands in stark contrast to participants from the rural (38.8%, CI: 31.9%-45.2%) and peri-urban (22.1%, CI: 16.1%-28.1%) participants who mostly had local interactions. Part-time and full-time employees from the urban community spent an overwhelming proportion of contact hours outside their community, 84.3% (CI: 78.3-89.5%) and 83.4% (77.9-88.1%), respectively. Participants from the peri-urban community spent the least amount of contact time outside the community (17.8%, CI: 11.1-25.2%).

## Discussion

By analysing social contact data from three geographically distinct communities in South Africa, we show a large degree of heterogeneity in contact patterns that could have substantial implications for the design and evaluation of TB interventions.

Our findings suggest that the efficacy of household contact tracing for TB (Context 1) may vary across different geographic settings. Consistent with other social contact and epidemiological studies, (12,27,28) participants across all three communities reported substantial contact hours at home. However, the proportion of contact varied considerably. While nearly three-quarters of contact hours occurred within homes in peri-urban (70.1%) and rural (76.2%) communities, less than half (46.1%) occurred within urban participants’ homes.

A potential explanation for the disparities in household contact hours is the exceptional difference in average household size. Urban participants lived with an average of 3.9 household members, compared to 15.2 and 10.9 in the peri-urban and rural communities, respectively. These large household sizes suggest that household contact tracing could be a particularly effective strategy for identifying and treating individuals with TB in these communities. Conversely, in the urban community where household sizes were much smaller, a substantial portion of contact occurred outside the home, highlighting the greater need for complementary strategies to detect people with TB. When designing and implementing screening programs for TB, it is important to consider the extent of household contact to optimise the effectiveness of these interventions.

We evaluated the differences in contact occurring outside the home, as it is relevant for infection prevention and control strategies in congregate settings (Context 2). Our results reinforce findings from other studies that public spaces play an important role in *Mtb* transmission.(4–6,14,29,30) Outside the home, urban participants had the highest proportion of contact hours in retail and office settings (19.2%), while rural participants had the highest proportion of contact hours in other people’s homes (25.1%). The highest proportion of contact hours reported by peri-urban participants occurred in community service buildings such as churches and clinics (19.7%). These findings demonstrate the diversity of contact patterns across different congregate settings, though further research is needed to explore the specific factors that may contribute to *Mtb* transmission in these places.

While contact hours in transport were relatively brief compared to other congregate settings, other studies have shown that overcrowding and poor ventilation in vehicles may increase the risk of *Mtb* transmission.(31–33) In the urban community, Deol et al. found that modes of transport generally had better ventilation compared to buildings.(31) This suggests, that for the urban community, transport may not carry as substantial an infection risk due to ventilation as might be expected. However, further research examining ventilation and transmission risk in transport across diverse community settings is needed to fully understand the role of transport in *Mtb* transmission.

The types of congregate settings visited varied substantially across communities, which underscores the importance of tailoring IPC measures to local contexts. While more expensive IPC methods such as germicidal UV lights and mechanical ventilation can be used in clinical settings,(25,34) more accessible approaches like opening windows,(31,35) and wearing masks (25,34) could also drastically reduce *Mtb* transmission. Prioritising accessible IPC efforts in high-contact settings may improve their impact on community-wide incidence. For example, encouraging open windows and/or promoting mask use in urban retail and office buildings, peri-urban community buildings, and rural homes could be promising strategies to reduce *Mtb* transmission. By combining social contact surveys with aerobiology and molecular data, future research could help identify the specific congregate settings where transmission is most likely to occur. This information may be used to target delivery of IPC interventions where they may have the greatest impact.

Considering each community as a cluster, we analysed contact occurring outside the community to assess mobility, a potential source of contamination in cluster randomised trials (Context 3). Mobility between clusters and the wider population can dilute the intervention impact, leading to a potential underestimation of the true effect.(16) Previous CRTs, such as ZAMSTAR and HPTN 071 (PopART), have reported high participant mobility as a limiting factor in demonstrating population-level reductions in TB burden.(36,37) Our social contact analysis reinforces this concern; urban participants spent nearly 70% of their contact hours outside the community. In contrast, rural and peri-urban participants spent 38.8% and 22.1%, respectively.

The observed disparities in contact hours outside the community underscore the importance of considering social contexts in CRTs. The high mobility of urban participants may be specific to this environment, with factors like employment opportunities and proximity to a major city potentially influencing contact and movement patterns. By integrating social contact surveys into trial design, researchers can identify the likely geographic range of mobility, and design clusters based on these patterns to mitigate contamination.(26) Furthermore, mathematical models can incorporate contact survey data to simulate various cluster selection and intervention delivery scenarios, enabling an optimised trial design before significant resources are invested.(26)

There are several limitations to our analysis. While this study provides valuable insights into the contact patterns of participants in these three communities, the findings may not be directly generalisable to other populations. Instead, our results highlight that substantial differences in contact patterns can exist between different communities in the same country. Furthermore, the survey methodology and the analysis presented here can serve as a framework for assessing contact patterns in other populations.

As with any survey, the social contact data collected were subject to measurement error and potential biases that may have impacted the reliability of the data. Participants may have had difficulty remembering and accurately reporting their interactions, including the number of people present, or the time spent in a building or in transport. Estimates of contact hours may also be less accurate when based on a random day in the past week, as was done in KwaZulu-Natal, compared to the day before the interview, as in Western Cape. The survey participants were mostly representative of each community’s age and sex distribution, except for higher participation among urban 18-19 year-olds and lower participation among peri-urban 30-39 year-olds. However, all analyses on contact hours were performed on the weighted dataset.

Another limitation was our approach to estimating contact hours. We capped the number of people present in buildings at 20 and in private cars at 10, with no other caps on modes of transport. Our sensitivity analyses with caps of 50 and 100 people in buildings show a modest effect on contact hours, however the distributions remain similar (Appendix; Table S3). Direct data were not available on whether contacts were members of the participant’s household or community, or whether they were residents of a different household or community. We therefore used the locations in which the contacts occurred as a proxy measure. Thus, we may have under- or over-estimated the proportion of contact hours that occurred between household members, and with people from outside the community. Although transport was excluded from the contact hour proportions outside the community (Figure 3B), due to the small proportion of contact in transport in congregate settings (Figure 4) this is unlikely to have had a large impact on the results.

## Conclusion

Our findings demonstrate the value of social contact data, and underscore the importance of tailoring interventions to specific community contexts, as a one-size-fits-all approach is insufficient to reduce TB. Household contact tracing may be most effective in the rural community where household contact hours were the highest. IPC measures are likely to have the greatest impact on transmission when implemented in congregate settings with high contact hours, and these settings varied by community. In the urban community, retail and office settings were the primary locations for interactions outside the home, compared to buildings providing community services and other people’s homes in peri-urban and rural communities, respectively. The risk of contamination in cluster randomised trials may be the greatest in the urban community due to the high mobility reported by urban participants. These findings show the value of social contact surveys and their application for developing locally informed interventions to reduce *Mtb* transmission. By leveraging this knowledge, policymakers and public health practitioners can develop more effective and equitable strategies to end TB in South Africa and other high burden settings.

## Data Availability

The social contact survey data used in this secondary analysis are available upon reasonable request to the survey principal investigator through an online repository: 1. McCreesh N, Middelkoop K, Mohlamonyane M. Cape Town social contact data [Internet]. London, United Kingdom: London School of Hygiene & Tropical Medicine 2022. https://doi.org/10.17037/DATA.00002756. 2. McCreesh N, Dlamini V, Edwards A, Olivier S, Dayi N, Dikgale K, et al. Social contact information from PIPSA residents in uMkhanyakude before and during the Covid-19 pandemic – data from the Umoya Omuhle and Covid Social Contacts studies [Internet]. Africa Health Research Institute (AHRI) 2021. https://doi.org/10.23664/AHRI.UOANDCSC.DATASET.2021 Upon publication, the code for this analysis will be available from: github.com/lekate15/SocialContact.

## Acknowledgements

We extend our gratitude to the social contact survey participants and to the fieldworkers who collected the data in Western Cape and KwaZulu-Natal. We thank the entire *Umoya omuhle* project team and have included a full list of members in the Appendix.

## Supporting information

**Table S1. Aggregated building groups based on survey response options.**

**Figure S1. Example of izigodi to catchment assignments in KwaZulu-Natal.** An example clinic catchment area is shown in yellow. The population centre of the red izigodi is located within the boundaries of the catchment area, therefore the izigodi is classified as within the community. While the blue izigodi intersects the catchment area, the population centre is outside, therefore the izigodi is classified as outside the community. This figure is for illustrative purposes only and does not represent boundaries from the KZN study communities.

**Figure S2. Decision tree for determining HIV status from survey responses.**

**Figure S3. Mean indoor contact hours per day occurring in buildings and transport by contact location and sociodemographic strata.** Contact in all congregate settings includes own home and transport; Contact outside the home includes transport; Contact outside the community excludes transport.

**Table S2. Observed survey population percentages compared to the survey sampling frame.**

**Table S3-1. Mean indoor contact hours in the urban community by sociodemographic strata.**

**Table S3-2. Mean indoor contact hours in the peri-urban community by sociodemographic strata.**

**Table S3-3. Mean indoor contact hours in the rural community by sociodemographic strata.**

**Table S4-1. Overall mean indoor contact hours in the urban community with person caps of 20, 50 and 100 in buildings and 10 in private cars, by sociodemographic strata.**

**Table S4-2. Overall mean indoor contact hours in the peri-urban community with person caps of 20, 50 and 100 in buildings and 10 in private cars, by sociodemographic strata.**

**Table S4-3. Overall mean indoor contact hours in the rural community with person caps of 20, 50 and 100 in buildings and 10 in private cars, by sociodemographic strata.**

**Table S5. Number and proportion of journeys based on origin and destination in among peri-urban and rural participants.**

**Table S6-1. Proportion of contact hours among urban participants occurring inside and outside the household and community.**

**Table S6-2. Proportion of contact hours among peri-urban participants occurring inside and outside the household and community.**

**Table S6-3. Proportion of contact hours among rural participants occurring inside and outside the household and community.**

**Table S6. Proportion of contact hours in congregate settings outside own home.**

## Statements

### Author contributions

**Conceptualization:** Kate E. LeGrand, Palwasha Khan, Nicky McCreesh

**Data curation:** Keren Middelkoop, Nicky McCreesh

**Formal analysis:** Kate E. LeGrand

**Investigation:** Kate E. LeGrand, Anita Edwards, Mbali Mohlamonyane, Njabulo Dayi, Stephen Olivier, Dickman Gareta, Robin Wood, Alison Grant, Richard White, Keren Middelkoop, Palwasha Khan, Nicky McCreesh

**Methodology:** Kate E. LeGrand, Palwasha Khan, Nicky McCreesh

**Supervision:** Palwasha Khan, Nicky McCreesh, Richard White

**Visualization:** Kate E. LeGrand

**Writing – original draft:** Kate E. LeGrand

**Writing – review & editing:** Kate E. LeGrand, Anita Edwards, Mbali Mohlamonyane, Njabulo Dayi, Stephen Olivier, Dickman Gareta, Robin Wood, Alison Grant, Richard White, Keren Middelkoop, Palwasha Khan, Nicky McCreesh

### Ethics approval

This study is a secondary analysis of social contact survey data collected from human participants. The survey protocol was approved by ethics committees at the London School of Hygiene and Tropical Medicine (LSHTM) (14520 & 14640), University of KwaZulu-Natal (UKZN) (BE662/17), and University of Cape Town (008/2018). Ethics approval for this secondary analysis was granted by LSHTM (28263) and UKZN (BREC/00005202/2023).

### Data availability statement

The social contact survey data used in this secondary analysis are available upon reasonable request to the survey principal investigator through an online repository:

1. McCreesh N, Middelkoop K, Mohlamonyane M. Cape Town social contact data [Internet]. London, United Kingdom: London School of Hygiene & Tropical Medicine; 2022. https://doi.org/10.17037/DATA.00002756.
2. McCreesh N, Dlamini V, Edwards A, Olivier S, Dayi N, Dikgale K, et al. Social contact information from PIPSA residents in uMkhanyakude before and during the Covid-19 pandemic – data from the Umoya Omuhle and Covid Social Contacts studies [Internet]. Africa Health Research Institute (AHRI); 2021. https://doi.org/10.23664/AHRI.UOANDCSC.DATASET.2021

Upon publication, the code for this analysis will be available from: github.com/lekate15/SocialContact.

